# Effect of CT-based material grouping on finite element strength and stiffness predictions in vertebrae with metastatic lesions

**DOI:** 10.64898/2026.08.20.26360953

**Authors:** Daniel Strack, Nils Rehtanz, Zahra Soltani, Mario Keko, Karupppasamy Subburaj, Ron N. Alkalay

## Abstract

**Introduction:** Metastatic spinal lesions substantially alter vertebral mechanical properties and increase fracture risk. Computed tomography (CT)-based finite element (FE) models can estimate vertebral strength, but their accuracy depends on how CT-derived material properties are represented. This study evaluated the effect of two material-grouping strategies on simulated strength and stiffness in metastatic vertebrae.

**Methods:** We compared Adaptive Clustering (AC) with Uniform fixed-width grouping in 44 vertebrae from 11 donors (8 osteolytic, 12 osteoblastic, 12 mixed, 12 no observed lesion (NOL)). FE models were generated based on CT scans with 2–500 material groups and compared for material-mapping error and simulated strength and stiffness. Overall and lesion-stratified agreement with experimental measurements was assessed in an exploratory analysis.

**Results:** AC showed significantly lower Young’s modulus root-mean-square error than Uniform (p < 0.05). Simulated strength and stiffness stabilised by 50 material groups. At 50 groups, simulated strength showed moderate correlation with experimental strength overall (R² = 0.57), strongest in NOL vertebrae (R² = 0.82) and lower in lesion-bearing vertebrae (R² = 0.41–0.59). Stiffness showed weaker correlation overall (R² = 0.27), highest in NOL vertebrae (R² = 0.48) and negligible in mixed lesions (R² = 0.007). Bland-Altman analyses indicated systematic underestimation of experimental fracture load.

**Discussion:** AC improved material-mapping fidelity, whereas increasing material groups beyond 50 had little influence on simulated strength or stiffness. Numerical stabilisation therefore did not imply experimental accuracy. Lesion-stratified findings were exploratory and should be interpreted cautiously because of limited subgroup sizes.

## 1 Introduction

Cancer is one of the leading causes of death [1]. Primary cancers can lead to metastasis growth in bone, with the spine being the most common site of bone metastasis, followed by the femur [2]. Bone metastases affect the bone microarchitecture, composition and cellular homeostasis [3], thereby degrading mechanical properties and increasing fracture risk [4]. Based on radiographic appearance, metastatic lesions are commonly classified as osteolytic, mixed or osteoblastic [5, 6]. When present in the spine, these lesions can compromise vertebral integrity and result in vertebral fractures, which cause pain, increase mortality and reduce quality of life [7, 8]. This clinical burden highlights the need to identify individuals at increased risk before fracture occurs. Thus, patient-specific fracture-risk assessment is critical for fracture prevention and preservation of patient mobility and survival.

Finite element (FE) analysis has been used extensively to assess the strength of healthy and osteoporotic bones [9, 10]. FE analysis has also been used to study the effects of metastatic lesions on bone strength [11], evaluate in vivo changes in patients with multiple myeloma [12], and simulate the effects of synthetic lesions [13]. Prior CT-based FE models of metastatic vertebrae have generally used material parameters derived from healthy bone or have tuned material parameters to match experimental vertebral behaviour under axial compression loading [11, 14, 15]. These approaches have yielded moderate to excellent correlations between experimental and simulated strength and stiffness, and have shown good agreement with internal strain distributions in both healthy and metastatic vertebrae [16]. However, these validation outcomes do not isolate the influence of specific modelling choices, such as how continuous CT-derived density and elastic modulus values are best discretised into a finite number of material groups to accurately represent bone tissue conditions. This lack of standardisation is particularly relevant in metastatic vertebrae, where osteolytic, osteoblastic, and mixed lesions substantially alter the distribution of density-based material properties [3, 4].

In CT-based FE models, continuous density-elasticity relationships are often discretised into a limited number of material groups to reduce computational cost and model complexity while retaining material heterogeneity. This discretisation may introduce mapping error that can propagate to predicted vertebral strength and stiffness. In a previous study, we demonstrated that both the grouping algorithm and the number of material groups influence association with experiments and computational efficiency in non-lesion vertebrae [17]. Metastatic lesions (osteolytic, osteoblastic, and mixed) present significant material heterogeneity [3, 4] that can substantially broaden and distort the element-wise distribution of density-based material properties. However, the impact of material grouping strategy and the number of material groups required to optimise the FE estimates of strength and stiffness has not been systematically evaluated in metastatic vertebrae spanning bone lesion phenotypes.

In this study, we investigated how discretising CT-based material properties affects vertebral FE predictions in a heterogeneous dataset of cadaveric vertebrae containing metastatic lesions. Specifically, we (i) compared Adaptive Clustering and Uniform Grouping using the root-mean-square error (RMSE) between the original CT-derived element-wise Young’s modulus field and the grouped material field, and used this RMSE to assess material mapping fidelity and convergence with an increasing number of material groups; (ii) quantified the effect of the number of material groups on predicted strength, as the primary endpoint, and stiffness; and (iii) performed an exploratory comparison with experimental measurements to assess the association and agreement between simulated and experimental outcomes overall and across lesion types. We hypothesised that compared to the uniform grouping approach, adaptive material grouping would reduce the number of material groups and associated modulus mapping error, thereby improving convergence of predicted strength and stiffness in metastatic vertebrae.

## 2 Methods

This study was approved by the institutional review board. A total of 44 vertebral specimens from 11 donors were analysed.

### 2.1 Specimen and image acquisition

We analysed 44 vertebral specimens obtained in a previous study [11] from 11 individual donors (3 female donors, mean age 60 years, range 49-71) harvested through the Anatomy Gifts Registry (Hanover, MD, USA). All donors had a history of a solid primary tumour (prostate, breast, esophageal, kidney or lung) [11].

#### I: Clinical CT imaging

Each spine was imaged with a clinical CT scanner (Aquilion 64, Canon Medical System (formerly Toshiba Medical), USA) using standard spine imaging protocol (125 kV, 60 ms, ROI: 8.0 in., matrix size: 512, slice thickness: 500 μm, 396 μm in-plane resolution and resampled to an isotropic resolution of 318.5 µm). Upon radiological review of the clinical CT (RNA) 44 vertebral specimens were reclassified as having osteolytic (8 vertebrae), osteoblastic (12 vertebrae), mixed (12 vertebrae) and no observed lesion (NOL, 12 that did not exhibit identifiable lesion on CT).

#### II. Mechanical specimen preparation

As previously detailed [11], the identified vertebral levels were extracted, cleaned and prepared for standardised mechanical testing. In short, the specimens were extracted and all soft tissue was removed. Subsequently, the pedicles were sectioned in the coronal plane proximal to the body to remove the posterior elements and a diamond-coated bandsaw (Exakt 300, Exakt Technologies, Inc., Germany) was used to section both endplates under constant water irrigation resulting in a plano-parallel vertebral body section.

#### III. Micro-CT imaging

Each plano-parallel vertebral body section was imaged in a micro-CT scanner (μCT100, Scanco Medical, Switzerland) yielding image data at 24.5 μm isotropic voxel size [11].

### 2.2 Experimental measurement

As previously detailed [11], each vertebra was compressed between two steel plates in a servo-hydraulic compression system (858 Mini Bionix II, MTS, Eden Prairie, USA) with the cranial plate mounted to the compression device with a ball joint. A uniaxial displacement was applied at a rate of 5 mm/min until failure was reached or the maximum force of the load cell (15 kN) was reached [11]. Strength was computed as the maximum compressive force attained on the load-displacement curve. Seven specimens reached the 15 kN load-cell limit before fracture occurred. For the exploratory linear regression and Bland-Altman analyses, these specimens were retained and the maximum recorded load of 15 kN was used as the experimental strength value. Stiffness was determined by linear regression over the 20–80% portion of the load-displacement curve prior to yield. Yield was identified using the 0.2% offset method.

### 2.3 Model preparation

Model preparation was achieved using the following stages.

#### Segmentation

The clinical CT scan and micro-CT scans were imported to 3D Slicer (www.slicer.org) [18]. A deep learning approach [19] was used to segment the vertebral levels from the clinical CT scan. For the micro-CT data, the “grow from seeds” algorithm available in 3D Slicer was used to segment each vertebral body, followed by Poisson’s smoothing operation. Using 3D Slicer rigid registration, each micro-CT segmented vertebral body volume was co-registered to the deep-learning (DL) segmented clinical CT vertebral body volume and a volume corresponding to the micro-CT volume extracted from the clinical CT volume. This operation resulted in a clinical CT vertebral volume corresponding to the volume of the experimental vertebra. An in-house-developed meshing tool [20] was used to obtain FE mesh models from the registered data using linear tetrahedral elements applying a target element size of 1 mm, as previously determined by Soltani et al. [15].

#### Material Definitions

Material parameters, including the Young’s modulus, were mapped based on grey-scale intensities using Bonemat [21] (V4 www.bonemat.org) and relationships listed in Table 1. Three material models were considered: (i) a linear-elastic isotropic model; (ii) a linear-elastic anisotropic model, in which the Young’s moduli in the x-and y-directions were scaled relative to the z-direction; and (iii) a nonlinear anisotropic elastic-plastic model incorporating yield and softening behaviour to determine simulated strength. For each vertebral specimen, a local coordinate system was manually defined, with the x-axis oriented anterior– posteriorly, the y-axis oriented medio-laterally and the z-axis defining the superior-inferior and compression direction. The anisotropic and nonlinear parameters of the model were automatically assigned using the PBMGA software tool [22].

**Table 1:** Parameters for the material model.

| Parameter | Relationship |
| --- | --- |
| Apparent density ($\rho_{app}$ in $Kg/m^3$) | $\rho_{app} = 5.1 + 0.7 \times HU$ |
| Ash density ($\rho_{ash}$ in $Kg/m^3$) | $\rho_{ash} = 0.6 \times \rho_{app}$; [23] |
| Young's modulus ($E$ in $MPa$) | $E_z = 4730 \times (\rho_{app})^{1.56}$; [24] |
| | $E_x = E_y = 0.333 \times E_z$; [25] |
| Poisson ratio ($V$) | $V_{xy} = 0.381$; [25] |
| | $V_{xz} = V_{yz} = 0.104$; [25] |
| Shear modulus ($G$ in $MPa$) | $G_{xy} = 0.121 \times E_z$; [25] |
| | $G_{xz} = G_{yz} = 0.157 \times E_z$; [25] |
| Maximum principal stress limit ($\sigma$ in $MPa$) | $\sigma = 137 \times \rho_{ash}^{1.88}, \rho_{ash} < 0.317$; [26] |
| | $\sigma = 114 \times \rho_{ash}^{1.72}, \rho_{ash} > 0.317$; [26] |
| Plastic strain ($\epsilon_{AB}$) | $\epsilon_{AB} = -0.00315 + 0.0728 \times \rho_{ash}$; [27] |
| Minimum principal stress limit ($\sigma_{min}$ in $MPa$) | $\sigma_{min} = 65.1 \times \rho_{ash}^{1.93}$; [27] |
| Plastic modulus ($E_p$ in $MPa$) | $E_p = -4000 \times \rho_{ash}^{2.05}$; [27] |

#### Material Grouping

Based on the variations in material distribution in metastatic vertebrae, we aimed to compare two methods of material grouping and study their impact on the stiffness and strength of metastatic human vertebral specimens. Following material application, each element was assigned a unique Young’s modulus. To reduce computational complexity, element-wise values were grouped using either Adaptive Clustering (AC) [17] or Uniform Grouping (Uniform) [22]. The Uniform Grouping algorithm calculates the minimum and maximum Young’s modulus values present in the specimen and based on the user-specified number of groups, defines material groups of equal size. For Adaptive Clustering, the algorithm employs a k-means-clustering based algorithm to group materials together. Details are described in the literature [17, 22]. We evaluated simulated results for both methodologies at 2, 5, 10, 20, 50, 100, 200 and 500 material groups.

#### Boundary conditions and solving

To reproduce the experimental loading conditions, the lower surface of each model was fully fixed and compressive loading was applied to the upper surface. Fracture load was estimated from nonlinear displacement-controlled simulations using an applied displacement of 0.5 mm, which was sufficient to capture fracture load in all models. Stiffness was calculated from linear-elastic load-controlled simulations using an applied compressive force of 3000 N. All models were solved using Abaqus (SIMULIA, Dassault Systems, version 2025) on a workstation computer (Z6, Intel Xeon W7, 32 cores, 128 GB RAM, Nvidia Quadro GV 100 32 GB computational accelerator, 2 TB PCIe-4.0 SSD drive, Hewlett-Packard, Palo Alto, California). The computational cost was measured as the time needed to execute the simulation, i.e. the solver time in Abaqus. Simulated results were extracted using an in-house Python script, and resulting force-displacement graphs (FDG) were processed to determine strength (defined as peak in the nonlinear simulation FDG) and stiffness (defined as slope in linear-elastic simulations FDG).

### 2.4 Statistical Analysis

All statistical analyses were performed in Python (version 3.11, https://www.python.org/) using the SciPy library (version 1.11.1, https://scipy.org/) [28].

We performed convergence analysis to assess the effect of material grouping on the distribution of the Young’s modulus E_z_. First, we computed the root-mean-square error as:

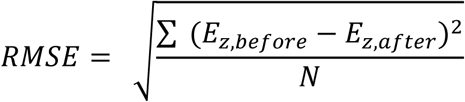

where *E_z_*_,*before*_ is the Young’s modulus before grouping and *E_z_*_,*after*_ is the Young’s modulus after grouping assessed for each element and *N* is the number of elements in a given bone. To determine the material-field convergence assessed with the Young’s modulus mapping RMSE, the number of material groups *k* was increased iteratively with convergence achieved when the relative change in RMSE compared with the value two steps earlier (*k* − 2) was lower than 1%. Using (*k* − 2) (rather than (*k* − 1)) avoided premature stopping caused by occasional one-step plateaus in RMSE. After convergence was achieved the RMSE values for 200 and 500 material groups were calculated for visualisation purposes.

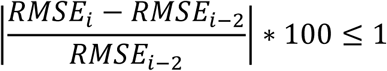

To visualise and analyse the stabilisation of strength values, we calculated the delta value in strength and stiffness for each specimen at each number of material groups k to the result at 500 material groups as:

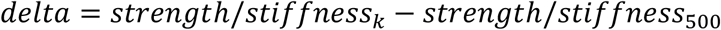

We used 500 material groups as the highest tested discretisation, consistent with prior work [29]. Moreover, we observed diminishing returns beyond ∼200 groups, with only marginal changes between 200 and 500 groups. Given the increased model complexity and computational cost associated with further increasing *k*, we did not test higher discretisation levels. Because RMSE evaluation does not require running Abaqus, we assessed material-field convergence for all *k* values in steps of one. FE analyses for global mechanical outcomes were performed only at predefined *k* values (2, 5, 10, 20, 50, 100, 200, 500), as simulating every *k* would be prohibitively time-consuming.

We used the Wilcoxon signed-rank test to compare Uniform and AC, with p < 0.05 considered statistically significant. To account for multiple specimens originating from the same donor, specimen-level values were first aggregated by calculating the median within each donor.

Linear regression analyses were performed at the vertebral level and did not account for within-donor clustering; therefore, the resulting R^2^ values were interpreted descriptively. Bland-Altman analysis was performed using the repeated-measures approach described by Bland and Altman [30] to account for multiple vertebrae originating from the same donor. Lesion-stratified analyses were considered exploratory because of the limited subgroup sample sizes and repeated observations within donors.

## 3 Results

Differences in material-property distributions among lesion types are shown for one exemplary specimen per lesion in Figure 1. To account for differences in mesh size, the distributions were normalised such that the area under each curve equalled one. The osteoblastic specimen showed a distribution shifted toward higher Young’s modulus values.

**Figure 1.**
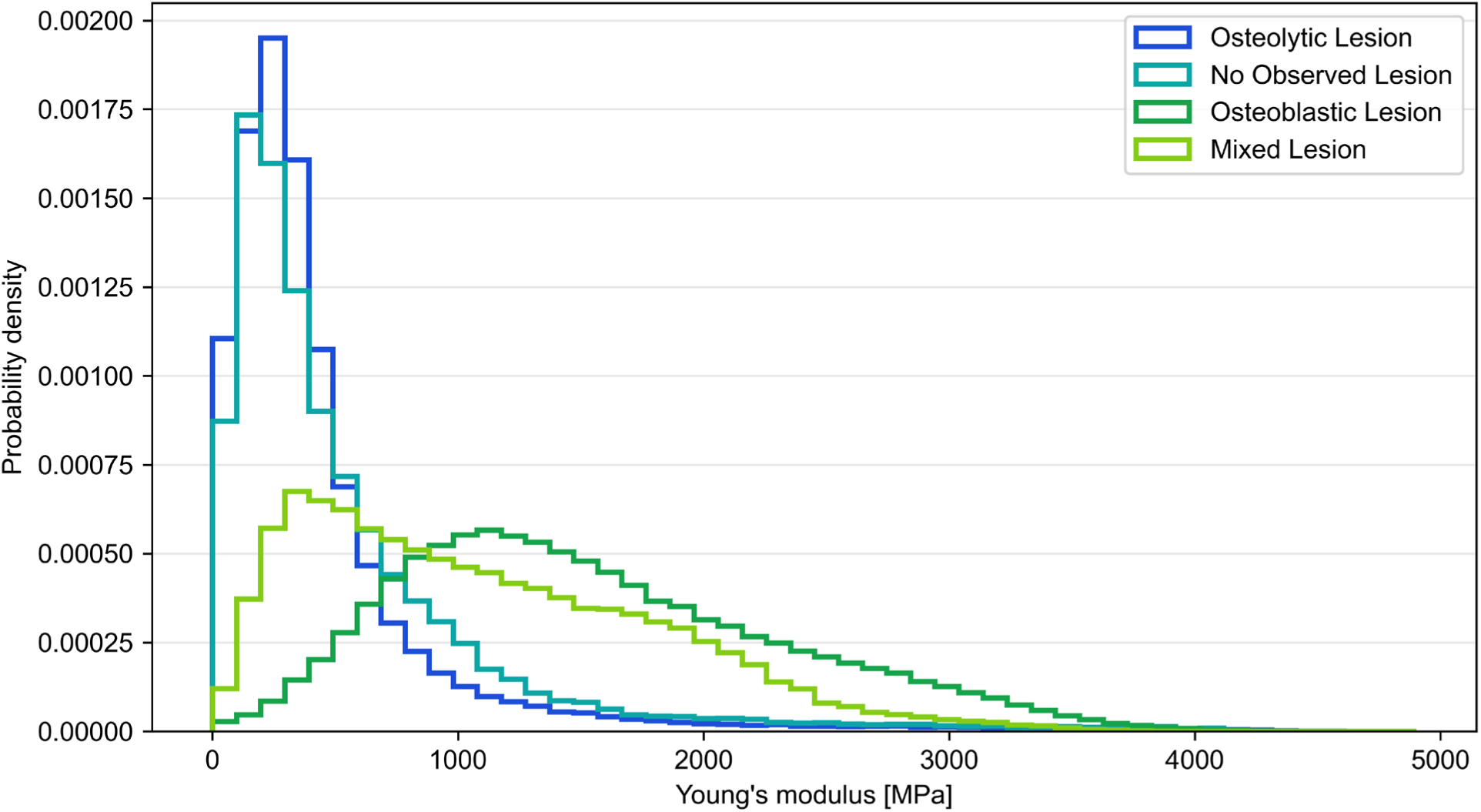
Normalised distributions of element-wise Young’s modulus values for one representative FE model from each lesion category (osteolytic, osteoblastic, mixed, and no observed lesion). Curves are probability densities (area under each curve equals 1) to account for different numbers of elements across models.

### 3.1 Impact of material grouping on RMSE

Figure 2 shows the convergence analysis results. The median number of material groups required for convergence (Figure 2A) is lower in the AC method across the overall dataset (138.5 vs. 140.5) and osteoblastic and osteolytic subgroups. In mixed and NOL subgroups the median values are nearly identical between Uniform and AC. Pairwise comparison between Uniform and AC using Wilcoxon signed-rank test showed no significant differences (p > 0.05).

**Figure 2.**
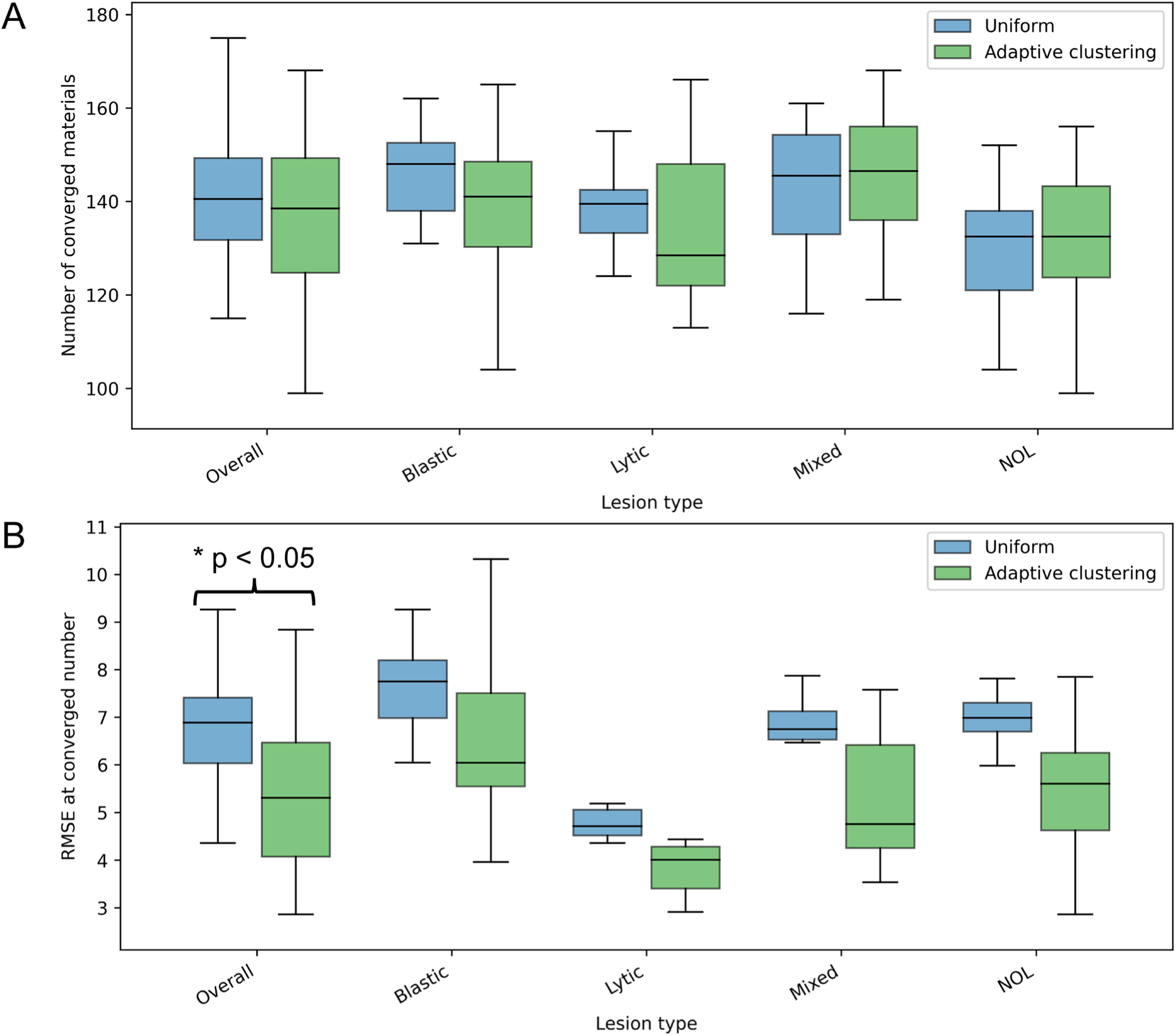
Box plot showing the number of materials at convergence (A) and the respective RMSE [MPa] (B). Wilcoxon signed-rank test showed significant difference when analysing the RMSE values in the overall dataset for AC and Uniform. Boxes indicate the median and interquartile range (IQR), whiskers extend to the most extreme observations within 1.5 times the IQR.

Figure 2B presents the results of the RMSE at the converged number of material groups. Across all vertebrae and in the subgroup analysis the median RMSE is lower in AC (5.31 MPa) compared to Uniform (6.89 MPa). Pairwise comparison showed significant differences in the overall dataset (p < 0.05).

### 3.2 Computational cost

We further explored the impact of material grouping on the computational cost. Figure 3 shows the median simulation time for all 44 specimens during the execution of the nonlinear simulation to assess FE-derived fracture load. Corresponding trends are observed for both AC and Uniform as with an increase of material groups the simulation time increases as well and an inflection point is observed between 20 and 200 material groups. We found a substantial reduction in simulation time when reducing the number of materials from 500 to 50 materials (−61% AC, −58% Uniform), and a further substantial reduction from 50 to 5 materials (−30% AC, −24% Uniform).

**Figure 3.**
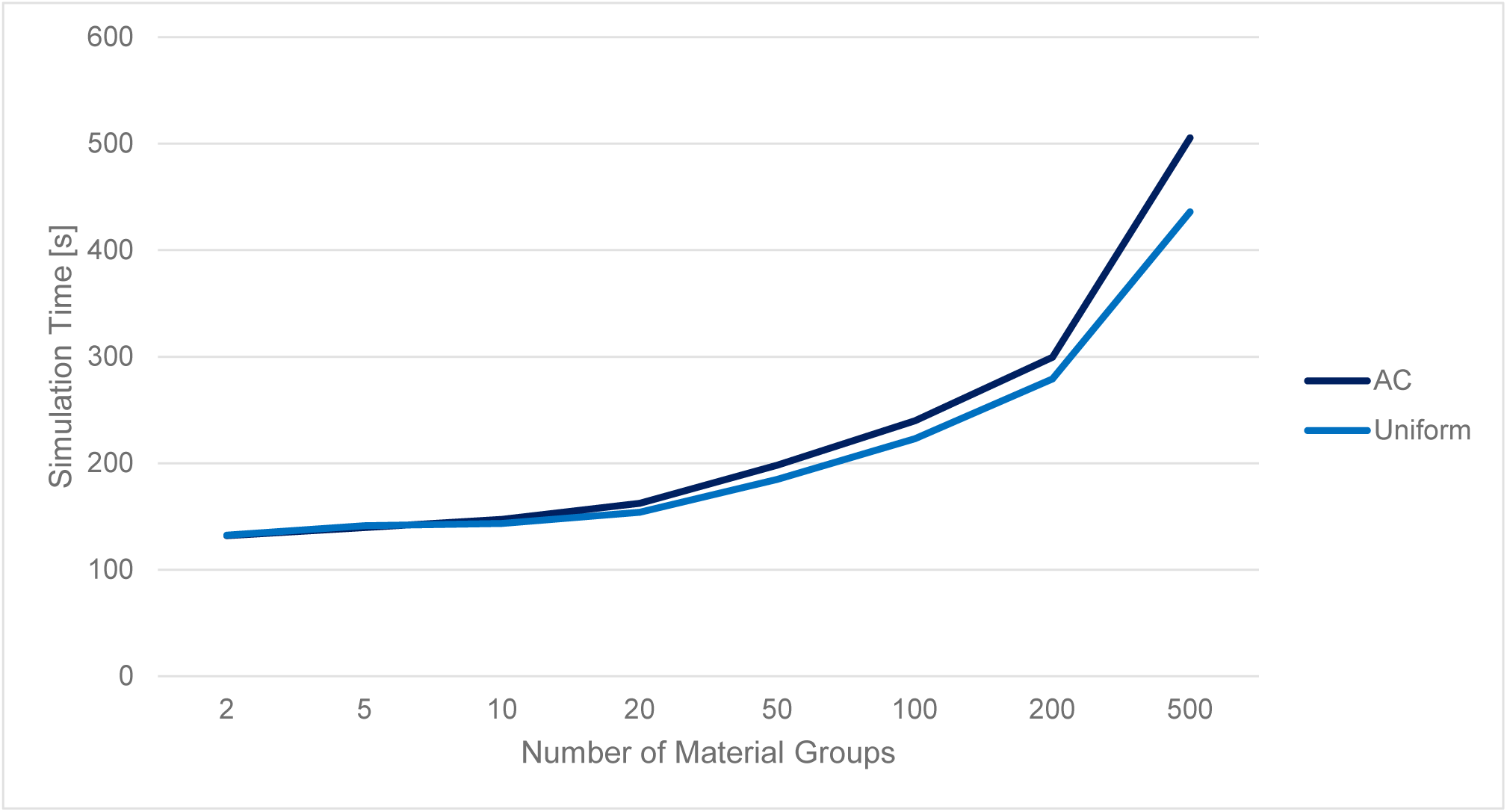
Median simulation time for Adaptive Clustering and Uniform Grouping

### 3.3 Impact of material grouping on strength

Figure 4 shows the stabilisation of simulated strength with increasing numbers of material groups. At two material groups the median value of Uniform was higher than of AC and the whiskers are smaller. At five and 10 material groups the median strength of Uniform was smaller than that of AC, but with similar whiskers. At 20 or more material groups, the medians and distributions were nearly identical between Uniform and AC.

**Figure 4.**
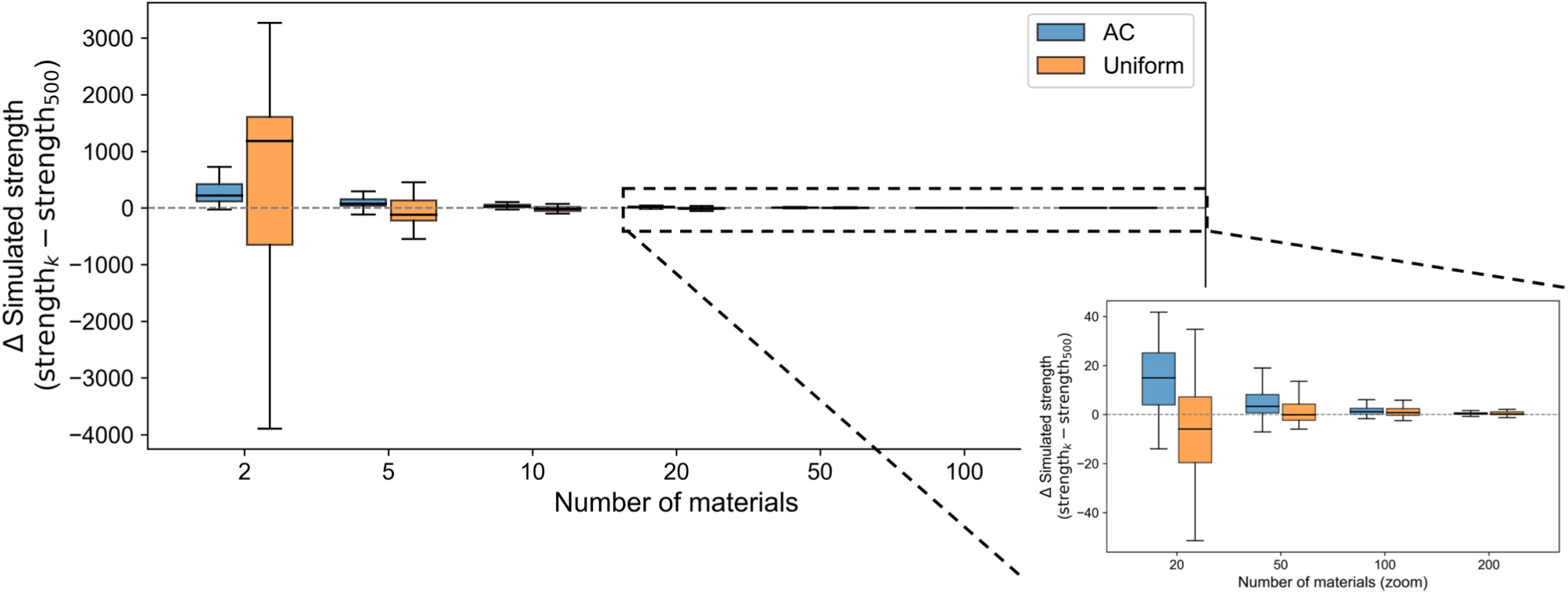
Difference in simulated strength (N) between each material-group count *k* and the 500-group reference. The inset shows results for 20–200 groups using an expanded y-axis

Because there was no visual improvement after 50 material groups, we calculated the delta of the strength value to 500 material groups seen in Figure 4. It can be observed that at 50 material groups the differences relative to the 500-group reference became small and showed little further change beyond 50 groups. At 50 material groups, the median difference relative to 500 groups was 3.52 N (0.12%) for AC and −0.13 N (−0.006%) for Uniform.

### 3.4 Exploratory lesion-stratified comparison between experiment and simulation

We compared the experimental results for strength with the AC simulated ones at 50 material groups. We observed an overall moderate correlation between experiment and simulated strength (R^2^ = 0.567, see Figure 5A) with a strong negative bias (mean difference: −5642.13N; 95% limits of agreement: −10898.03N to −386.24N) in the Bland-Altman plot (Figure 5B). Visual inspection of the Bland-Altman plot showed increasingly negative differences at higher fracture loads, suggesting possible proportional bias. Excluding the seven load-cell-limited specimens did not materially alter the overall interpretation; results are provided in Supplementary Material S6.

**Figure 5.**
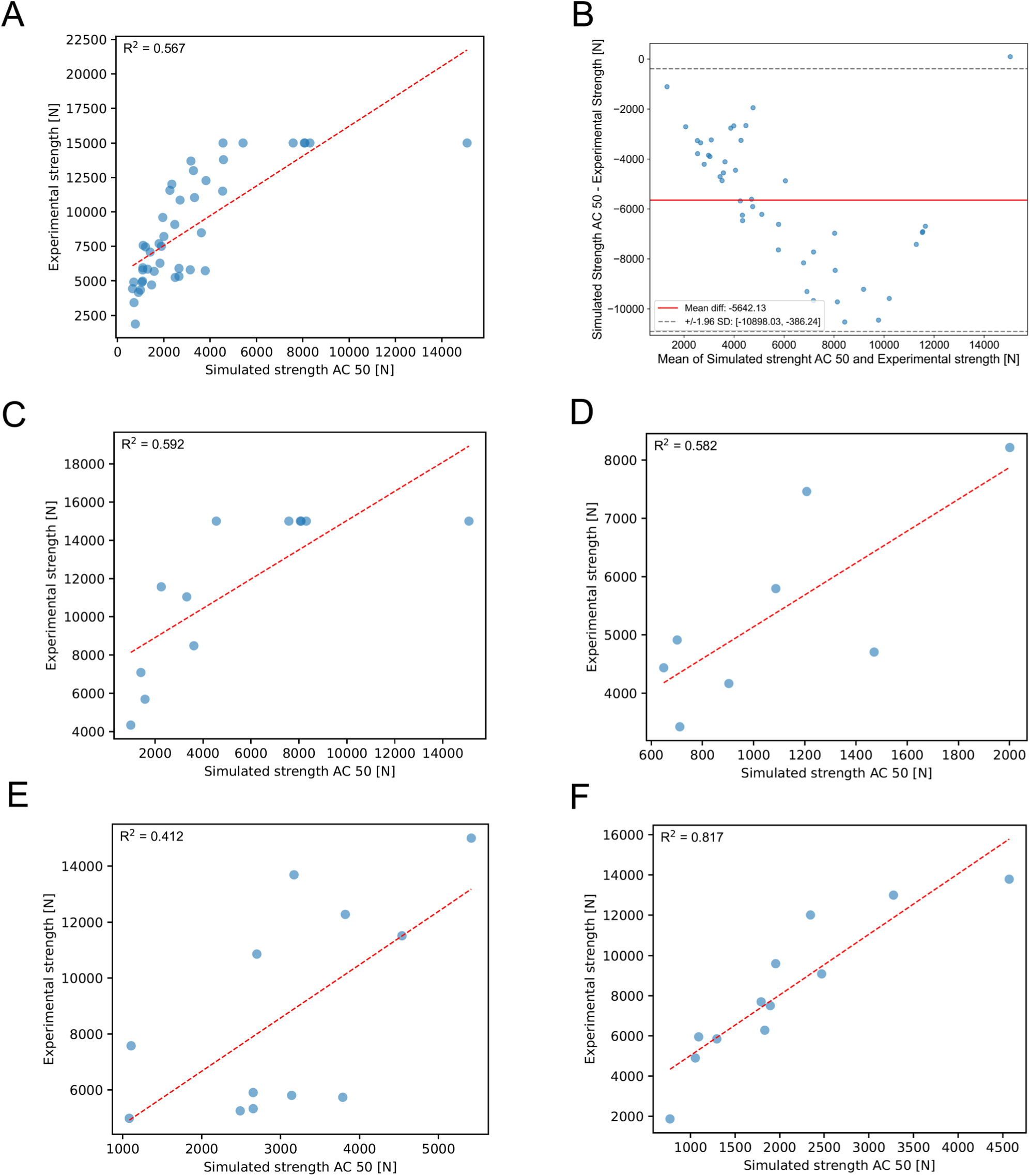
Bland-Altman and lesion-stratified correlation analysis between AC with 50 material groups and the experimental strength. (A) Correlation analysis pooled over the complete dataset. (B) Bland-Altman plot pooled over the complete dataset. (C) Correlation analysis stratified for vertebrae with osteoblastic lesions. (D) Correlation analysis stratified for vertebrae with osteolytic lesions. (E) Correlation analysis stratified for vertebrae with mixed lesions. (F) Correlation analysis stratified for vertebrae with NOL

After stratifying the correlation analysis for lesion type similar results for osteoblastic (R^2^ = 0.592, see Figure 5C) and osteolytic lesions (R^2^ = 0.582, see Figure 5D) were found. Correlation was slightly reduced in vertebrae with mixed lesions (R^2^ = 0.412, see Figure 5E). In contrast, a higher correlation was observed in vertebrae with NOL (R^2^ = 0.817, see Figure 5F).

### 3.5 Impact of material grouping on stiffness

Consistent with the strength result, data was collected and analysed for stiffness values obtained from the linear-elastic isotropic and anisotropic material model. Results closely followed the trend observed for simulated strength. Both AC and Uniform showed similar stabilisation behaviour, with only minor changes beyond 50 material groups (Supplementary Material Figures S1 and S3).

At high numbers of material groups, residual differences relative to the 500-group reference were numerically larger for AC than for Uniform. Conversely, at 2 and 5 material groups, Uniform showed larger differences and greater variability (Supplementary Material Figures S2 and S4).

The association between simulated and experimental stiffness was substantially weaker than that observed for strength (R^2^ = 0.269 Anisotropic, R^2^ = 0.259 Isotropic). Exploratory lesion-stratified analyses showed a similar pattern to strength, with the highest association observed in NOL vertebrae and the lowest in mixed lesions (See Supplementary Material Table S5).

## 4 Discussion

In this study, we quantified how discretising CT-derived material properties into a limited number of material groups affects FE predictions in vertebrae with metastatic lesions. We found that convergence of element-wise material-field fidelity (Young’s modulus mapping RMSE) required substantially more groups than stabilisation of global mechanical outputs: RMSE continued to improve up to approximately 140 groups (median across specimens), whereas simulated strength and stiffness stabilised by approximately 50 groups. At the convergence criterion Adaptive Clustering yielded a significantly lower RMSE than Uniform Grouping in the overall dataset, partially supporting our hypothesis. The same pattern was observed when considering subgroups defined by the lesion type; however, the differences were not statistically significant, likely reflecting the smaller sample sizes within the subgroups. Further, strength and stiffness differences between algorithms were most pronounced at very low numbers of groups (e.g., 2–5). In exploratory validation against experiments, association was reduced in vertebrae affected by metastases, particularly mixed lesions, which may indicate that a material model developed on healthy bone may not fully capture metastatic alterations.

Adaptive Clustering converged with a lower number of material groups in the overall dataset compared to Uniform (median number of groups 138.5 vs. 140.5), however this difference was not statistically significant (p>0.05). Similar trends were observed in the lesion-stratified subanalysis, in osteoblastic and osteolytic lesions the median number of converged groups is lower in AC compared to Uniform, while the number is nearly identical in mixed and NOL subgroups. Additionally, we report the remaining RMSE at the number of material groups required for convergence and found that there is a significant difference in the overall group, with AC having the lower RMSE. Thus, AC showed greater material-mapping fidelity: the median number of groups required for convergence was numerically lower, whereas the remaining discretisation error was significantly lower than with Uniform Grouping. To the best of our knowledge, this is the first study to systematically analyse the material discretisation error introduced by material grouping. Previous studies have either evaluated the changes in average Young’s modulus or simulation output metrics [17, 31–33] or used a fixed number of material groups [29, 34].

It was observed that the strength values for both AC and Uniform stabilised by 50 material groups and yielded nearly identical results. In contrast, at a reduced number of material groups (2, 5) substantial differences between the methods were present (Figure 4). At 2 material groups Uniform overestimated the strength compared with the 500-group reference value and at 5 slightly underestimated it, whereas AC gradually approached the 500-group reference value (Figure 4). We further observed a consistent increase of computational cost with increasing number of materials. Comparing the computational cost (Figure 3) at 500 to 50 materials we observed a more than half reduced simulation time (−61% AC, −58% Uniform), and a further substantial reduction from 50 to 5 materials (−30% AC, −24% Uniform). While for several applications a number of 50 material groups is manageable and the choice of grouping algorithm therefore may play a subordinate role, in resource-constrained environments like highly dynamic crash simulations [35] the choice of grouping algorithm may be consequential.

Because no compelling differences in strength predictions were observed beyond 50 material groups and differences between AC and Uniform were marginal, the correlation analysis to experimental results was carried out using AC and 50 material groups. We found moderate correlation with experimental strength in the overall dataset (R^2^ = 0.567) for AC with 50 groups. This is substantially lower than the observed correlation of Soltani et al. [15] in 10 metastatic specimens, where they achieved R^2^ = 0.99 when tuning the material parameters to match the experiments and high correlation of R^2^ = 0.83 when using a global set of calibrated parameters over all 10 specimens. This is comparable to the results obtained by Stadelmann and colleagues [11] who used a material model derived from healthy bone and analysed 45 metastatic specimens achieving a good correlation (R^2^ = 0.78), hinting that metastatic material calibration improves strength predictions. Follet and colleagues [14] used a yield criterion similar to that applied in our study and reported a moderate correlation (R^2^ = 0.68) for the same experimental dataset as Stadelmann [11]. The higher correlations reported in some of these studies suggest that differences in constitutive formulation and material calibration may influence predictive performance; however, cross-study differences prevent attribution of the higher correlations to a single modelling choice. The substantial negative bias (−5642.13 N) observed in the Bland–Altman analysis (Figure 5B) indicates systematic underestimation of experimental strength by the simulations. This underestimation may partly reflect limitations of the nonlinear material model, which was derived from healthy bone, and was not materially improved by increasing the number of material groups beyond approximately 50. Similarly, Follet et al. [14] reported a substantial negative bias of −3749 N using a comparable modelling approach in metastatic vertebrae.

Exploratory, lesion-stratified analyses suggested reduced correlation between simulated and experimental strength in vertebrae affected by metastases compared with vertebrae with no observed lesions (NOL). In descriptive subgroup plots, the NOL group showed the highest apparent association (R² = 0.817), whereas the metastatic subgroups showed lower associations (osteoblastic R² = 0.592; osteolytic R² = 0.582; mixed R² = 0.412). These lesion-stratified values should be interpreted cautiously because specimens were not independent (multiple vertebrae per donor) and donor representation differed across lesion categories; thus, the stratified correlations are intended as hypothesis-generating rather than definitive between-group comparisons. Our observations differ from Stadelmann and colleagues [11], who found that mixed lesions had the highest association (R^2^ = 0.90) compared to osteoblastic and osteolytic (R^2^ = 0.64 and R^2^ = 0.76), acknowledging the exploratory nature of their analysis because of the limited sample size within lesion-stratified analysis. Furthermore, Garavelli et al. [16] reported comparable correlation between digital volume correlation (DVC)-measured and FE-simulated displacements in healthy versus metastatic specimens (R² = 0.69 vs. 0.64), emphasising that the apparent impact of metastasis may depend on the endpoint studied and on model assumptions and boundary conditions.

Similar results regarding stabilisation of strength were observed for stiffness. Regardless of modelling bone as linear-elastic isotropic or anisotropic, stiffness predictions stabilised at about 50 material groups. Anisotropic and isotropic setups also showed comparable stabilisation behaviour for Uniform grouping with initially (2 material groups) overestimating the stiffness of 500 material groups, followed by a gradual approach to the 500-group reference from below (5-20 material groups). Analysing the correlation with experimental results we found an overall moderate to low correlation (anisotropic R^2^ = 0.269; isotropic R^2^ = 0.259), substantially reduced from the values observed for strength (R^2^ = 0.567). Introducing anisotropic material behaviour did not alter the overall correlation. This observation is consistent with Krone and Schuster [36] who found that material anisotropy was not needed to describe femoral bone tissue in elastic bending. These values are also considerably lower than the observed values by Soltani et al. [15] (R^2^ = 0.9). However, Garavelli reported correlations between experimentally recorded reaction force prior to failure and FE reaction forces as R^2^ = 0.71, highlighting that this reduced to R^2^ = 0.2 if one value of high force is excluded [16]. Exploratory lesion-stratified analysis disclosed similar patterns as in strength. Numerically highest correlation was found for NOL (anisotropic R^2^ = 0.480; isotropic R^2^ = 0.473), whereas the mixed-lesion subgroup showed a near-zero association (anisotropic R^2^ = 0.007; isotropic R^2^ = 0.004). Osteoblastic lesions had comparable correlations to the overall dataset (anisotropic R^2^ = 0.245; isotropic R^2^ = 0.232) with osteolytic lesions being closer to NOL (anisotropic R^2^ = 0.463; isotropic R^2^ = 0.373). As above, these subgroup findings should be interpreted cautiously due to repeated measures within donors and small subgroup sizes. The near absence of correlation for mixed lesions is notable and may reflect complex load-sharing in highly heterogeneous vertebrae affected by both osteolytic and osteoblastic compartments, which may not contribute proportionally to compressive load transfer under the applied boundary conditions [37, 38]. More broadly, the clinical value of stiffness as an endpoint in vertebral fracture risk assessment remains less established than strength, and many studies focus primarily on predicted failure load [39–41]; in this context, stiffness is best viewed as a secondary endpoint and as motivation for future work evaluating whether stiffness adds independent information beyond strength in metastatic vertebrae.

### Limitations

Several limitations should be considered when interpreting our results. The primary aim of this study was to evaluate material grouping, whereas lesion-stratified analyses were exploratory. We acknowledge strong negative bias in Bland-Altman plots pointing to a systematic underestimation of the experimental results. The causes of this substantial negative bias are likely multifactorial and cannot be determined from the present study. One potential contributor is the use of a yield criterion derived from femoral rather than vertebral bone [26]. In future work, the material model, particularly the parameters defining yield strength, should be recalibrated and independently validated. We carried out donor-adjusted Wilcoxon signed-rank test to account for several vertebral specimens coming from the same donor. Overall and lesion-stratified R^2^ values are therefore presented for descriptive purposes; given the repeated-measures structure (multiple vertebrae per donor) and limited subgroup sizes, we did not pursue additional donor-aware subgroup association metrics. We therefore refer to this analysis as hypothesis-generating throughout the manuscript, acknowledging this inherent limitation. Mechanical testing of bones remains extremely costly, reducing the amount of available experimental data for lesion-stratified analysis. A recent study by Bruno et al. [42] showed that 3D-printed vertebrae, including metastases, based on medical images could serve as a cost-reducing alternative mirroring behaviour of donor bones. Furthermore, seven specimens reached the load-cell limit of 15 kN before fracture occurred, indicating that their true fracture loads exceeded the measurable range. For the exploratory linear regression and Bland-Altman analyses, these specimens were retained and assigned the maximum recorded load of 15 kN. Treating these censored observations as exact values may have affected the estimated associations and agreement. Notably, six of the seven specimens belonged to the osteoblastic subgroup and may therefore have disproportionately influenced the lesion-stratified results. We nevertheless retained these specimens to avoid further reducing the already limited sample size. Sensitivity analyses excluding the specimens that reached the load-cell limit, including the correlation and Bland-Altman analyses, are provided in Supplementary Material S6. Consequently, the corresponding regression and Bland-Altman results should be interpreted with caution.

## 5 Conclusion

In this study we found that Adaptive Clustering produced significantly lower material-field discretisation error, although the number of groups required to meet the material-field convergence criterion did not differ significantly between grouping approaches. Secondly, despite continued reductions in material-field error beyond 50 groups, global strength and stiffness predictions stabilised by approximately 50 material groups and were nearly identical between grouping approaches. At low material group counts before stabilisation of strength and stiffness, Adaptive Clustering showed smaller deviations from the 500-group reference than Uniform Grouping. This might be important in resource-constrained dynamic simulation where the number of material groups needs to be low. If a higher number of material groups (e.g., 50) is tolerable, the choice between AC and Uniform has only a marginal effect on global mechanical predictions. Importantly, numerical stabilisation did not imply experimental accuracy: simulated fracture strength was systematically underestimated, while stiffness showed only weak association with experimental measurements.

## Declaration of generative AI and AI-assisted technologies in the manuscript preparation process

During the preparation of this work, the authors used ChatGPT for language proofing. The authors reviewed and edited the output as needed and take full responsibility for the content of the published article.

## Author Contributions

Conceptualization: DS, RNA Methodology: DS, NR, MK Formal analysis & Investigation: DS Visualization: DS, Writing – Original Draft: DS, RNA; Writing – Review & editing: DS, NR, ZS, MK, KS, RNA Supervision & Funding acquisition: RNA, KS

## Data Availability

All data produced in the present study are available upon reasonable request to the authors

## Acknowledgements

This research was supported by the Aarhus University Research Foundation under the grant AUFF-E-2022-7-12 (Karupppasamy Subburaj). The National Institute of Arthritis and Musculoskeletal and Skin Diseases supported the work of R. Alkalay under its Research Project Grants (AR075964).

